# Improved Sensitivity For Detection Of Clinical Deterioration When Diagnostic Pathology And Patient Trends Are Included In Machine Learning Models

**DOI:** 10.1101/2024.10.20.24315403

**Authors:** Jonathan D. Greenberg, Leo C.E. Huberts, Angus Ritchie, Sze-Yuan Ooi, Gordon M. Flynn, Graeme K. Hart, Blanca Gallego

**Author notes:** **Primary contact:** Blanca Gallego. **Secondary contact:** Jonathan Daniel Greenberg. **Declarations of interest:** None.

## Abstract

**Objectives:** This study aimed to develop and validate a machine learning model to predict deterioration using Australian hospital data, paying particular attention to the role of predictors not included in current scoring systems.

**Design:** Retrospective cohort study using electronic health records from a large metropolitan health service.

**Setting:** General hospital wards, excluding the Emergency Department, Intensive Care Unit, or Palliative Care.

**Participants:** Inpatients over the age of 18.

**Main Outcome Measures:** The primary outcomes of deterioration were mortality and ICU transfer within 24 hours of a newly available observation. A Gradient Boosted Tree model was estimated using patient demographics, vital signs, pathology results, and linear trends. Resulting feature importance was investigated using Shapley values. The model performance was validated against existing scoring systems, including Between the Flags (BTF) and the Modified / National Early Warning Score (MEWS/NEWS).

**Results:** A Gradient Boosted Tree was developed from 121,608 patients and tested in 20,605 patients. The model, named aWARE, demonstrated higher discriminative ability (AUROC_mortality_=0.93, AUROC_ICU transfer_=0.84), and calibration when compared to baseline scores. Overall, the 10 most influential features unique between both outcomes were age, oxygen saturation to inspired oxygen ratio, respiratory rate, white cell count, venous lactate, heart rate to systolic blood pressure ratio, albumin, oxygen saturation, urea and heart rate. Of these, only 3 are included in BTF.

**Conclusion:** The machine learning model proposed in this study identified more deteriorating patients and produced less false positive alerts than Between the Flags. Feature importance highlighted the deficit between strong predictors of deterioration and the parameters used in current scoring systems.

## INTRODUCTION

Early Warning Systems (EWS), or Rapid Response Systems (RRS), were developed over 30 years ago to identify patient deterioration before an adverse event is observed. In New South Wales, the ‘Between the Flags’ (BTF) system triggers clinical reviews (‘yellow zone’) or Medical Emergency Team (MET) calls (‘red zone’) based on derangement of vital signs^1,2^.

Concerned staff may also trigger emergency responses and the calling criteria can be manually modified to allow personalisation of the rapid response or account for the patient resuscitation status or end of life wishes.

EWS implementation has reduced cardiac arrests, in-hospital mortality, and Intensive Care Unit (ICU) lengths of stay, yet limitations of the almost 15-year-old BTF system remain^3–5^. The BTF heuristic trigger rules rely on a small set of selected vital signs, leaving other relevant information (such as pathology findings) to be considered at the discretion of individual clinicians. Trigger rules do not account for natural vital sign fluctuation nor the interrelation between multiple patient variables^1^. Numerous false positive alerts lead to alarm fatigue and a reluctance to escalate, and yet many deteriorating patients are left undetected, with approximately 43% of unplanned admissions to ICU in Australia in 2020 occurring without an antecedent MET call^6, 7^.

The Modified Early Warning Score (MEWS) and the National Early Warning Score (NEWS) are aggregated scoring systems used internationally based on a weighted linear combination of selected vital sign parameters^8–10^. While this may provide a more accurate assessment of deterioration than BTF, it is also hindered by non-adherence as manual calculation is required^11^.

### Machine Learning Prediction of Deterioration Risk

The incorporation of machine learning and electronic monitoring systems may provide an avenue to reduce patient adverse outcomes^12,13, 14^. Although not exhaustive, Table 1 is a representation of promising and relatively simple machine learning algorithms of in-hospital deterioration across the world.

**Table 1:** Comparison of current prominent machine learning models of deterioration in ward-based hospital patients.

| Author | Year | Country | Tool Name | Model | Cohort | Outcomes | Predictors | Performance |
| --- | --- | --- | --- | --- | --- | --- | --- | --- |
| Escobar, G. et. al.<sup>19</sup> | 2012 | USA | *Advanced Alert Monitor (AAM)* | Logistic regression | 102,422 adult ward admissions across 14 hospitals, 2006-2009 | Transfer to ICU, in-hospital mortality | Patient demographics, time of day, length of stay, physiological derangement score, comorbidity score, vital signs laboratory results | Composite outcome: AUROC = 0.775 |
| Rothman, M. et. al.<sup>20</sup> | 2013 | USA | *Rothman Index* | Logistic regression | 22,265 adult patients at one hospital in one year (2004). Paediatric, obstetric, psychiatric presentations excluded | In-hospital mortality | Vital signs, nursing assessments, laboratory results, cardiac rhythm | AUROC = 0.93 when predicting 24h mortality |
| Loekito, E. et. al.<sup>21</sup> | 2013 | Australia | *LabMet* | Logistic regression | 71,453 Emergency Department patients, 2000-2006 | MET calls, ICU admission, death within 24h or following calendar day | Laboratory results (9 variables), patient demographics | AUROC = 0.69 (MET calls), 0.82 (ICU admission), 0.90 (death) |
| Churpek, M. et. al.<sup>22</sup> | 2014 | USA | *Electronic Cardiac Arrest Triage (eCART)* | Logistic regression | 59,301 adult patients at one hospital, 2008-2011 | Cardiac arrest or transfer to ICU | Vital signs, demographics, laboratory values, length of stay | AUROC = 0.87 for cardiac arrest, AUROC = 0.76 for transfer to ICU |
| Bell, D. et. al.<sup>23</sup> | 2021 | Australia | *Deterioration Index* | Logistic regression | 258,732 adult patient admissions, 2 private hospitals, 2016-2019 | In-hospital mortality, transfer to ICU, urgent surgery, rapid response alert | Patient demographics, vital signs, laboratory results, linear trends of observations | Composite outcome: AUROC = 0.89 |
| Pimentel, M. et. al.<sup>17</sup> | 2021 | United Kingdom | *Hospital-Wide Alerts Via Electronic Noticeboard (HAVEN)* | Gradient Boosted Tree | 496,710 patient (>16y) admissions over 4 hospitals, 2012-2017 | Cardiac arrest, transfer to ICU | Patient demographics, comorbidities, vital signs, laboratory results, observation variability | Composite outcome: AUROC = 0.901 |
| Romero-Brufau, S. et. al.<sup>18</sup> | 2021 | USA | *Mayo Clinical Early Warning Score (MC-EWS)* | Gradient Boosted Tree | 104,240 adult patients across 2 hospitals, 2010-2011 | Cardiac arrest, rapid response call, transfer to ICU | Patient demographics, vital signs, laboratory results, nursing assessments | Composite outcome: AUROC = 0.932 |

ICU, Intensive Care Unit; AUROC, Area under Receiver Operating Characteristic Curve

Gradient Boosted Trees are a tree-based learning method which may better capture the complex medical data interactions than linear models, can handle missing data, and are more efficient computationally than higher order neural networks^15, 16^. Recently developed models of patient deterioration have shown successful performance with gradient-boosted trees^17, 18^.

### Study Aims

This study aims to: 1) develop and validate a simple but efficient machine learning model to identify deterioration in ward patients using electronic medical records (eMR) data from New South Wales (NSW), Australia; 2) compare the predictive performance of this algorithm against other proposed available tools; and 3) analyse and quantify the value added by predictors not currently included in implemented early warning systems in NSW.

## METHODS

### Data

This study used a data set of routinely collected electronic medical data from emergency, inpatient and outpatient hospital visits in a large metropolitan multicentre health system between January 2019 and July 2021.

### Study population

The study population consisted of a retrospective cohort of in-hospital admissions by patients over the age of 18. Admissions were included from a variety of in-patient services, including medical, surgical, obstetric and psychiatry. Admissions located in the Emergency Department, Intensive Care Unit, and palliative care wards, or those with no recorded measurements, were excluded. An illustration of this cohort design is shown in the Supplementary Material (Figure S1).

### Primary Outcomes

The primary outcomes for prediction were in-hospital mortality and unplanned transfer to ICU in the 24 hours following any newly available patient data. Unplanned transfer to ICU was defined as any transfer from an included hospital ward to the ICU, while transfers directly from operating theatres were typically considered planned and therefore excluded^24^.

Coronary Care Units (CCU) or High Dependency Units (HDU) were included as wards in this analysis, and hence transfer to these locations was not considered under the outcome of unplanned transfer to ICU.

### Predictors

Patient data, including age, gender, observation time, vital signs, select pathology results from full blood count, electrolytes urea creatinine, liver function tests, and venous blood gas analyses were extracted. Variables were chosen based on literature review of deterioration factors, and frequency within the dataset. A full list of these variables can be found in the Supplementary Material (Figure S2). Fraction of inspired oxygen was estimated using the method of delivery and oxygen flow rate^25^. Three clinically used interaction predictors were constructed: the ratio of oxygen saturation to estimated fraction of inspired oxygen (P/F Ratio), the ratio of urea to creatinine, and the ratio of heart rate to systolic blood pressure (Shock Index), having shown previous feature importance in deterioration modeling^17^. To capture patient trajectory and improve model performance, three linear trends of each feature were constructed^23, 26^. These were the gradients of variable change over time from the baseline first measurement of that variable after admission, from the previous measurement, and from the measurement two prior.

### Algorithm

A Gradient Boosted Tree algorithm was used to make predictions every time a new vital sign or pathology result was made available in the eMR. We called it the ‘ai-driven WArning and REsponse’ (aWARE) algorithm. Where no value was available for specific measurements, the values were imputed directly from the previous reading. Since death and ICU admission are competing outcomes, we chose to model these two outcomes separately, rather than as a composite outcome. Fig. 1 is a visual representation of how the model integrates predictors in real time to continuously update deterioration risk as new patient data is collected.

**Figure 1.**
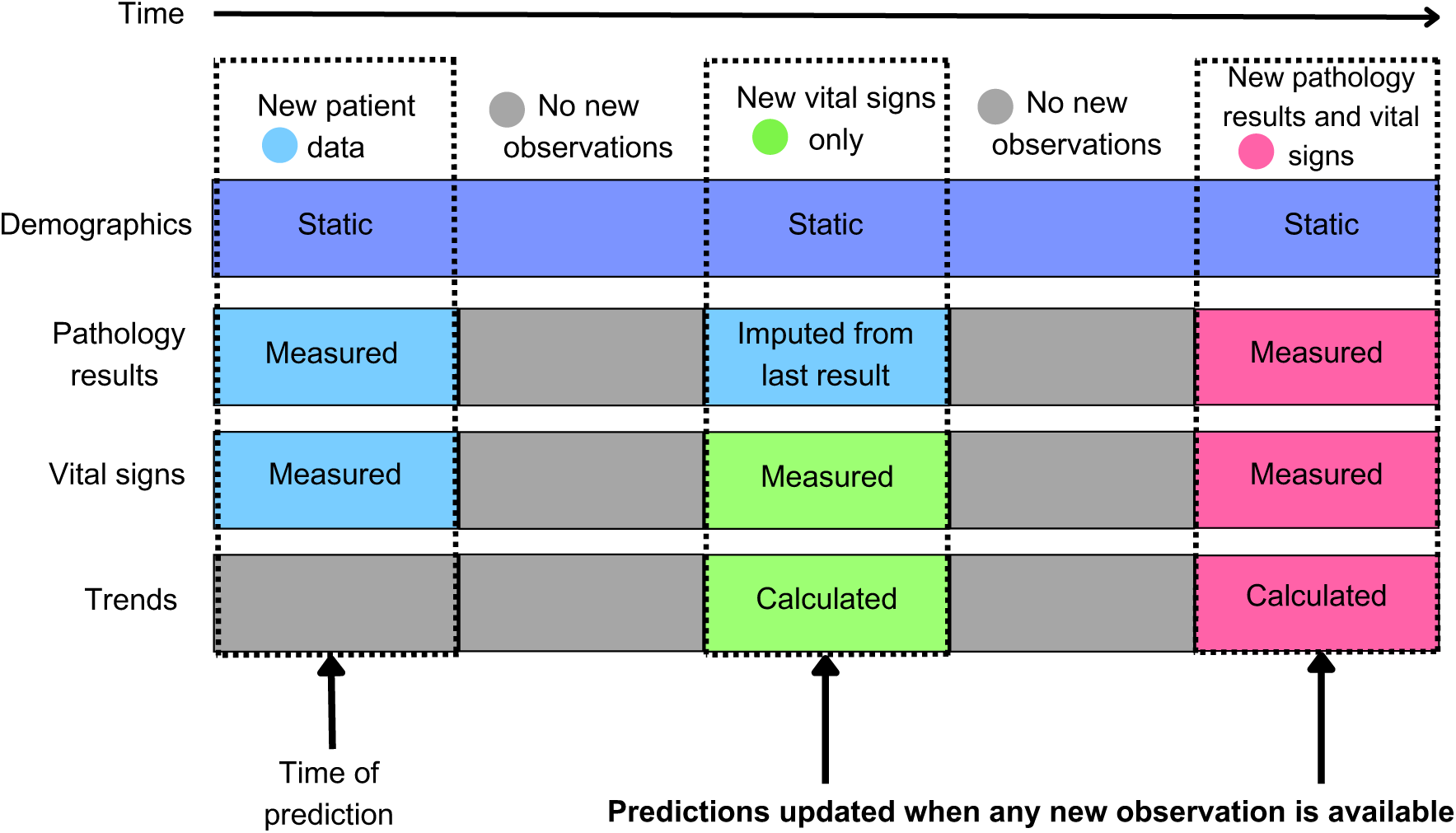
Schematic of model predictions updates. Time progresses from left to right. New colours represent new data input, either measured or calculated. Vertical dashed boxes represent the time of new predictions in accordance with new patient data input.

### Model development and validation

The data was split into training (70%), validation (10%) and test (20%) sets. A 5-fold cross-validated F1-score selected the best model hyperparameters, and isotonic regression calibrated probabilities. The final model was evaluated in the test set using predictive performance measures of discrimination (precision, recall, area under receiver operating characteristic curve (AUROC)) and calibration (Brier Score).

### Comparisons with other proposed algorithms

As data pertaining to actual observed alerts in the study period was not available, BTF alerts were calculated by simple conditions of calling criteria. MEWS and NEWS scores were calculated for each observation set. A logistic regression model was developed analogous to the eCART tool for comparison^22^. The Deterioration Index, HAVEN model and MC-EWS models, among others presented in Table 1, were not able to be directly compared due to missing predictor or outcome data. Using the holdout test set, the performance of the aWARE was compared against BTF, MEWS, NEWS and eCART with the same outcome of in-hospital mortality and unplanned ICU transfer in under 24 hours.

### Software

The analysis was conducted in Python (v3.9.13) using the SKLearn (v1.0.2) and XGBoost (v2.0.0) libraries

### Ethics approval

Ethics approval for this study was provided by the Sydney Local Health District Human Research Ethics Committee under the project ‘Data derived Risk assessment using the Electronic Medical record through Application of Machine Learning’ (DREAM). Local reference number: CH62/6/2018-203. REGIS reference number: 2019/PID09922.

## RESULTS

### Descriptive Statistics

Of 228,520 hospital admissions extracted from the DREAM database, 103,021 ward patients, across 154,654 admissions, were included in the study. Demographics of patients split by primary deterioration outcomes are compared in Supplementary Material (Table S1). There were 5,780,061 sets of patient results, of which 46,929 (0.81%) occurred within 24 hours of a deterioration event. The final model contained 125 predictors: 33 unique patient variables and 92 corresponding trends (Supplementary Material Table S2).

### Performance Metrics

Calibration curves (Fig. 2(a,b)) display how closely the calibrated model probability predictions match observed event proportions. aWARE showed better calibration than the other scores, with the strongest Brier scores of 0.0013 and 0.0068 for mortality and ICU transfer, respectively (lower scores indicating better accuracy). aWARE outperformed all baseline scoring systems in discriminative ability for both outcomes (Table 2). AUPRC was a comparatively small number across all scores, owing to the imbalanced prevalence of deteriorating patient observation points (0.81%). The Receiver Operating Characteristic curves (Fig. 2(c,d)) visualise discriminative performance across prediction thresholds.

**Figure 2.**
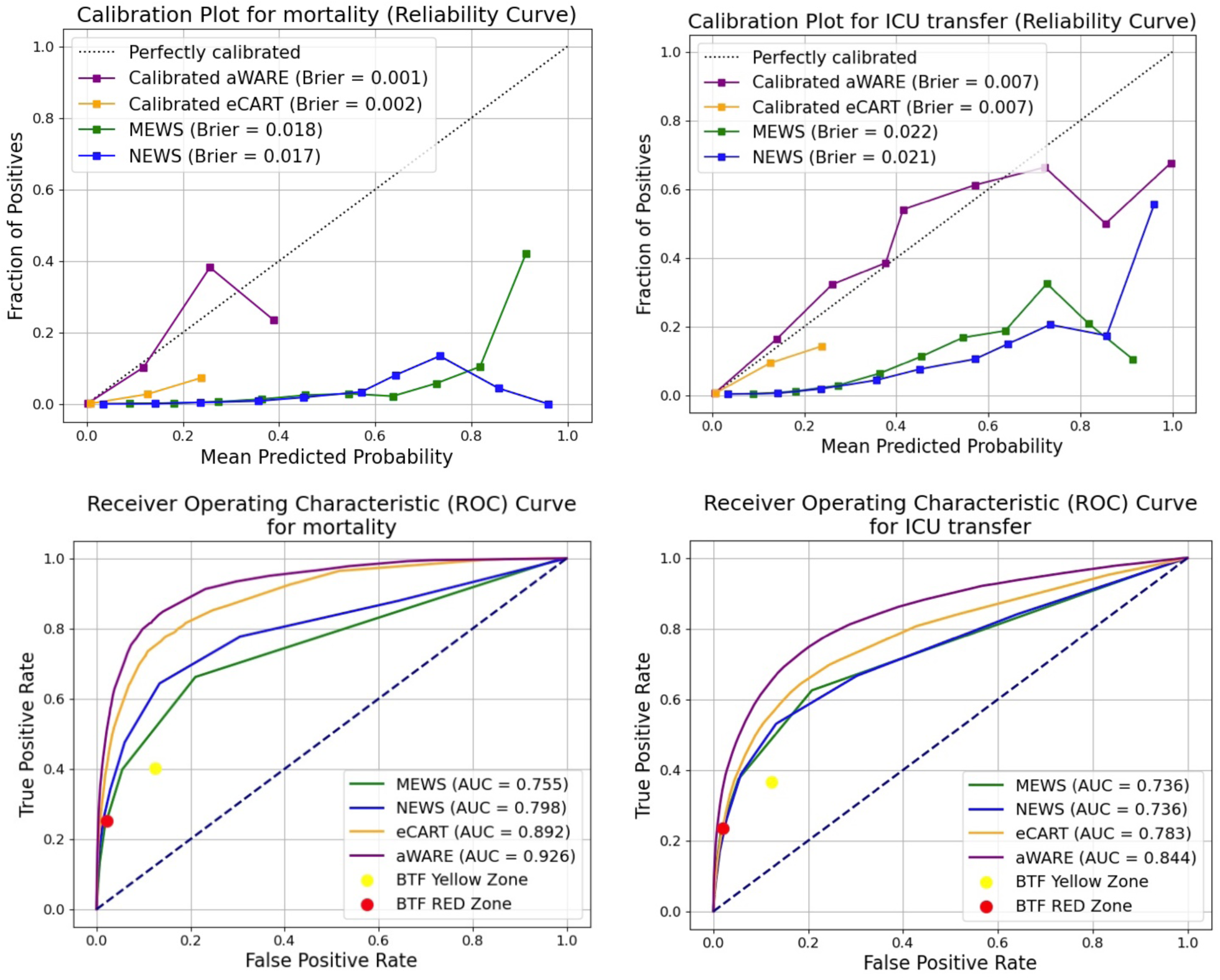
(a)(top left): Calibration curve for all scores predicting mortality. (b)(top right): Calibration curve for all scores predicting unplanned ICU transfer within 24h. (c)(bottom left): Receiver Operating Characteristic Curve for all scores predicting mortality in the entire test set. (d)(bottom right): Receiver Operating Characteristic Curve for all scores predicting ICU transfer in the entire test set. Brier score is shown for each model in calibration plots. Between the Flags Yellow Zone and Between the Flags Red Zone are represented by a singular point of predictive performance on Receiver Operating Characteristic curves, as these are not continuously modelled scores. aWARE, ai-driven WArning and REsponse system; BTF, Between the Flags; MEWS, Modified Early Warning Score; NEWS, National Early Warning Score; eCART, Electronic Cardiac Arrest Triage score; AUC, Area under Curve.

**Table 2:** Comparison of discriminative performance metrics in the entire test set for all scoring systems across primary prediction outcomes.

| **Prediction of mortality** |  |  |
| --- | --- | --- |
| **Scoring System** | **AUROC (95% CI)** | **AUPRC (95% CI)** |
| BTF Yellow Zone | - | - |
| BTF Red Zone | - | - |
| MEWS | 0.755 (0.742 – 0.768) | 0.012 (0.012 – 0.021) |
| NEWS | 0.798 (0.785 – 0.812) | 0.019 (0.018 – 0.026) |
| eCART | 0.892 (0.884 – 0.901) | 0.028 (0.073 – 0.092) |
| aWARE | 0.926 (0.920 – 0.933) | 0.064 (0.060 – 0.082) |
| **Prediction of ICU transfer** |  |  |
| **Scoring System** | **AUROC (95% CI)** | **AUPRC (95% CI)** |
| BTF Yellow Zone | - | - |
| BTF Red Zone | - | - |
| MEWS | 0.736 (0.729 – 0.741) | 0.047 (0.057 – 0.065) |
| NEWS | 0.736 (0.730 – 0.742) | 0.040 (0.044 – 0.050) |
| eCART | 0.783 (0.778 – 0.789) | 0.050 (0.066 – 0.074) |
| aWARE | 0.844 (0.839 – 0.845) | 0.124 (0.124 – 0.139) |

Between the Flags is not able to be modelled continuously and rather represents a singular threshold of prediction. BTF, Between the Flags; MEWS, Modified Early Warning Score; NEWS, National Early Warning Score; eCART, Electronic Cardiac Arrest Triage score; aWARE, ai-driven WARNING and REsponse system, AUROC, Area under Receiver Operating Characteristic Curve; AUPRC, Area under Precision-Recall Curve

### Model Performance matched to Between the Flag Alert Zones

Where a true positive is defined by a calculated alert or model prediction followed by mortality or ICU transfer within 24 hours, metrics of sensitivity and specificity may be calculated for BTF. For aWARE, the threshold at which a positive prediction is made was adjusted to match the closest specificity of each BTF Zone calling criteria, constructing two potential model ‘alert’ zones (aWARE Yellow and aWARE Red) that simulate current escalation practises (Table 3) and do not increase risk of false positive predictions. For each of these threshold zones, and for both outcomes of prediction, aWARE outperformed the sensitivity and precision of BTF, such that aWARE statistically only increases true positive predictions. Supplementary Material Figures S2&3 show the percentage of deteriorating patients identified by BTF and aWARE Red preceding primary outcome.

**Table 3:**
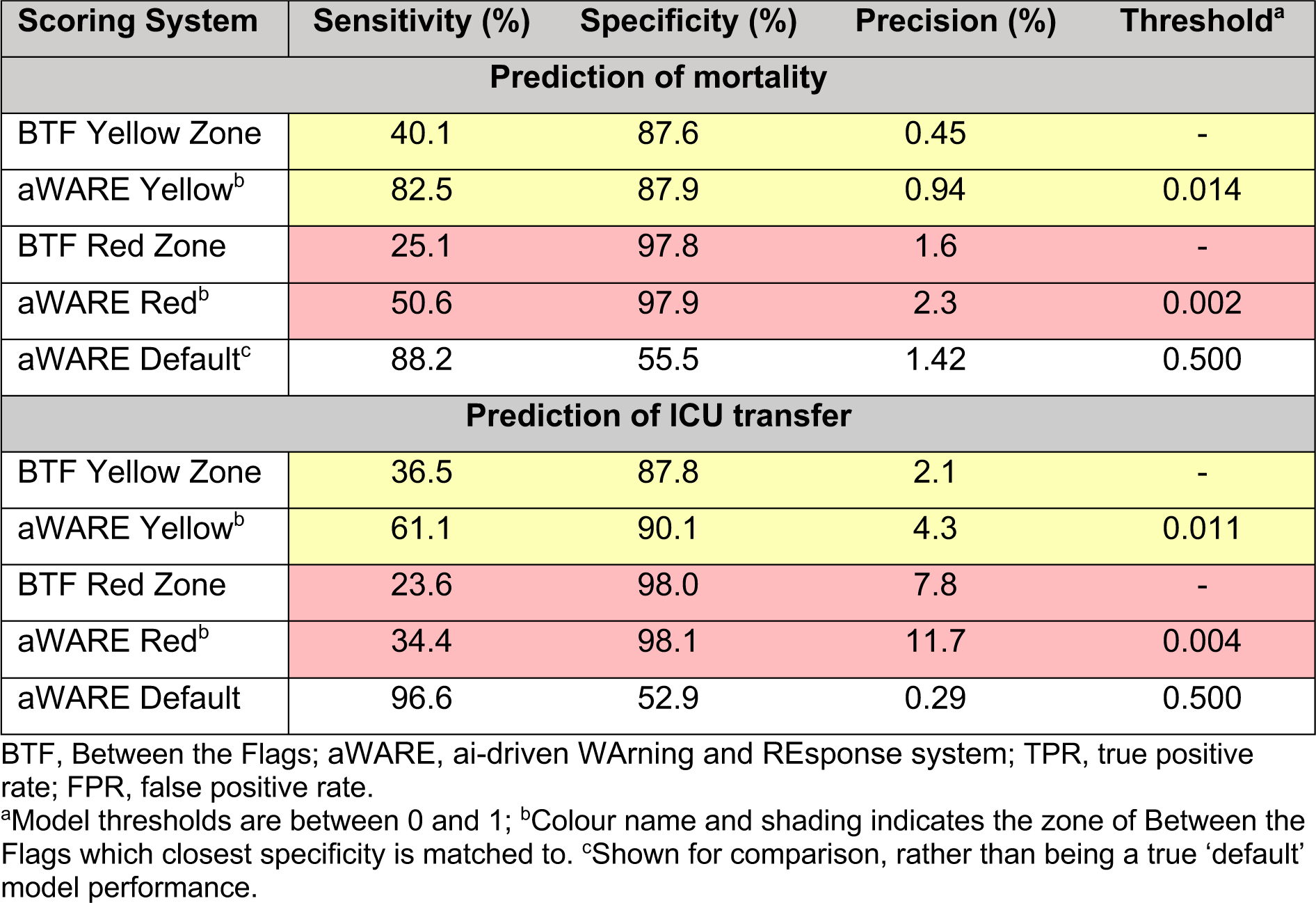
Comparison of performance metrics predicting mortality and ICU transfer in entire test set for Between the Flags and aWARE with varying thresholds.

### Feature Importance

We calculated Shapley values for each feature to explain their contribution to the model’s predictions, considering all possible feature combinations^27^. Figure 3 shows the top 10 model predictors ranked according to their relative importance as estimated by their mean absolute Shapley values. The 10 most influential features unique between both outcomes were age, SpO2:eFiO2, respiratory rate, white cell count, venous lactate, Shock Index, albumin, oxygen saturation, urea and heart rate. Of these, only three are included in BTF, while four were pathology results, two were vital sign interaction variables, and one was demographic.

**Figure 3.**
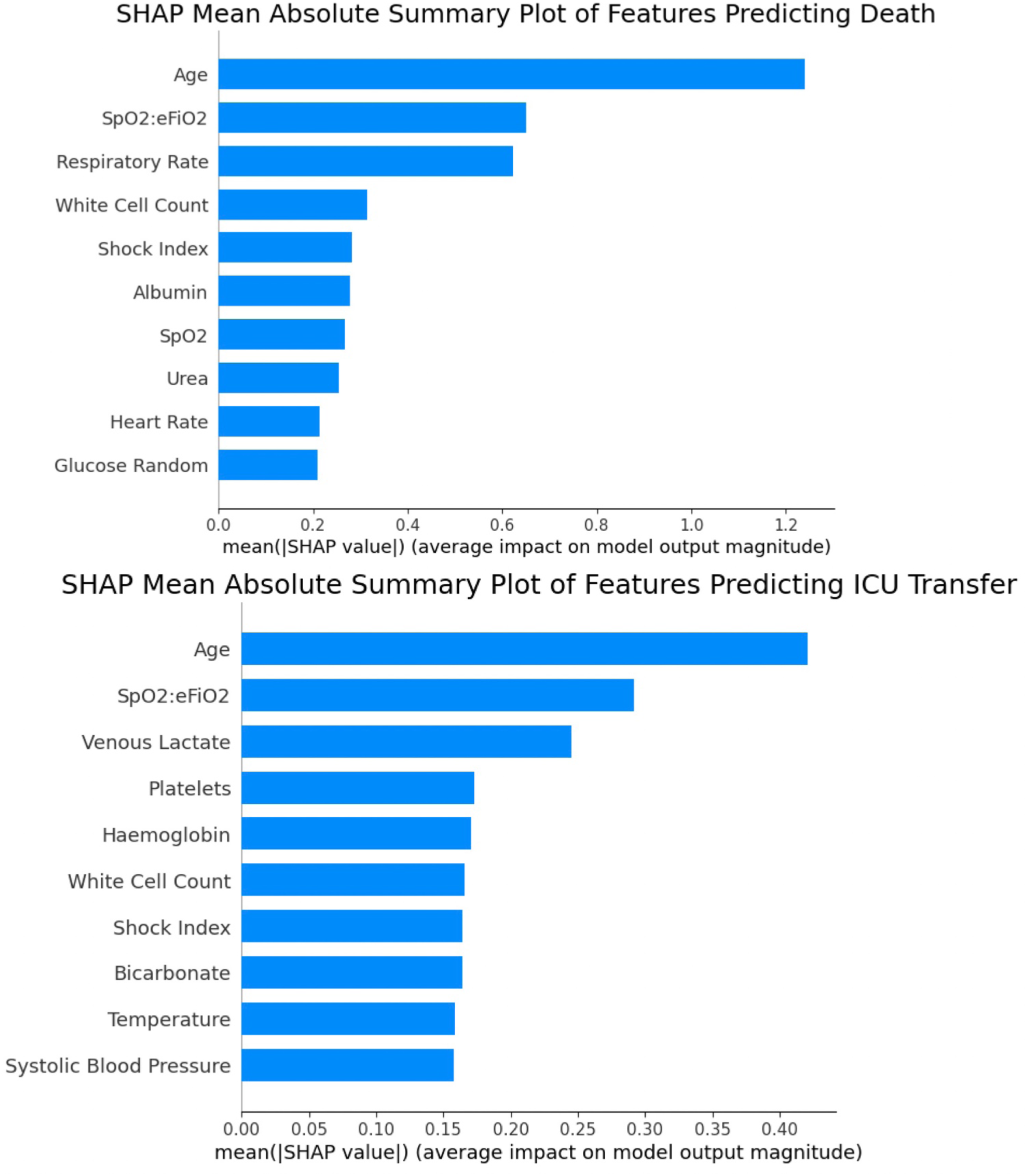
(a)(top): Mean absolute Shapley values of model features which predict mortality. Figure 3(b)(bottom): Mean absolute Shapley values of model features which predict ICU transfer. SpO2:eFiO2, ratio of oxygen saturation to estimate fraction of inspired oxygen; Shock Index, ratio of heart rate to systolic blood pressure

Adding demographics and pathology findings, interaction terms, and trends stepwise increases the predictive performance for mortatility and ICU transfer (Table 4).

**Table 4:**
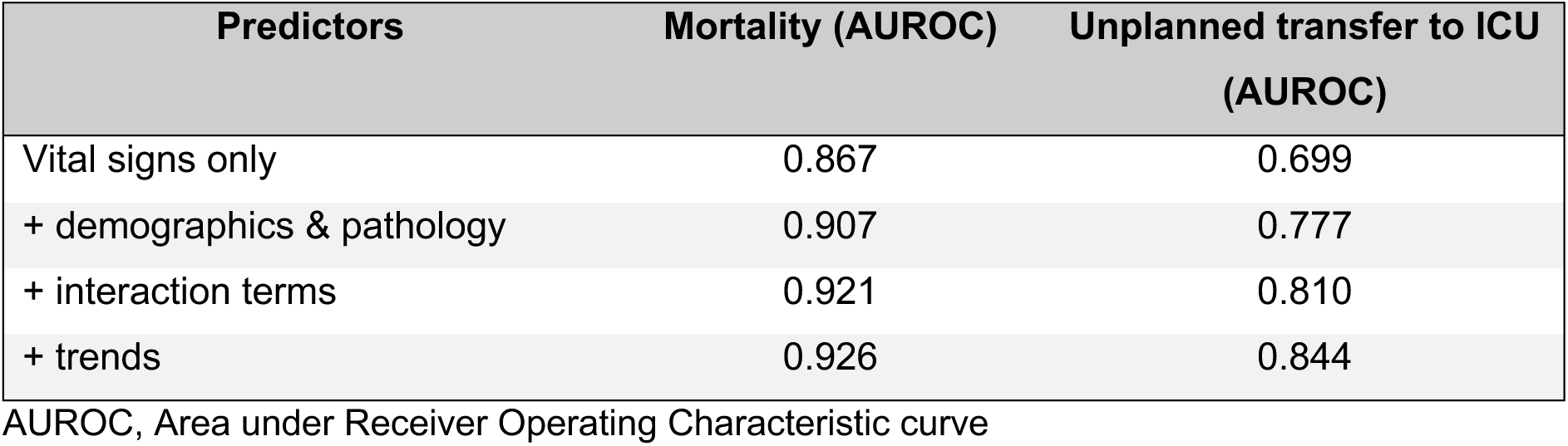
Model performance as patient predictors are added.

Shapley partial dependence plots (Figures 4&5) for the top four predictors of each outcome illustrate in detail how feature value influences the model’s predictions. Figure 4 shows that older age (>85), lower SpO2:eFiO2 (<4.5), higher respiratory rate (>20 breaths per minute) and white cell counts above the normal range are strong predictors of short-term death. Figure 5 indicates that the risk of ICU transfer was higher for patients with lower SpO2:eFiO2, higher venous lactate and haemoglobin outside the normal range. The probability of ICU transfer became lower for the youngest (<45) and the oldest (>85) age cohorts. Figures S4 and S5 in the Supplementary Material provide an overview of how prediction varies with other top predictors.

**Figure 4:**
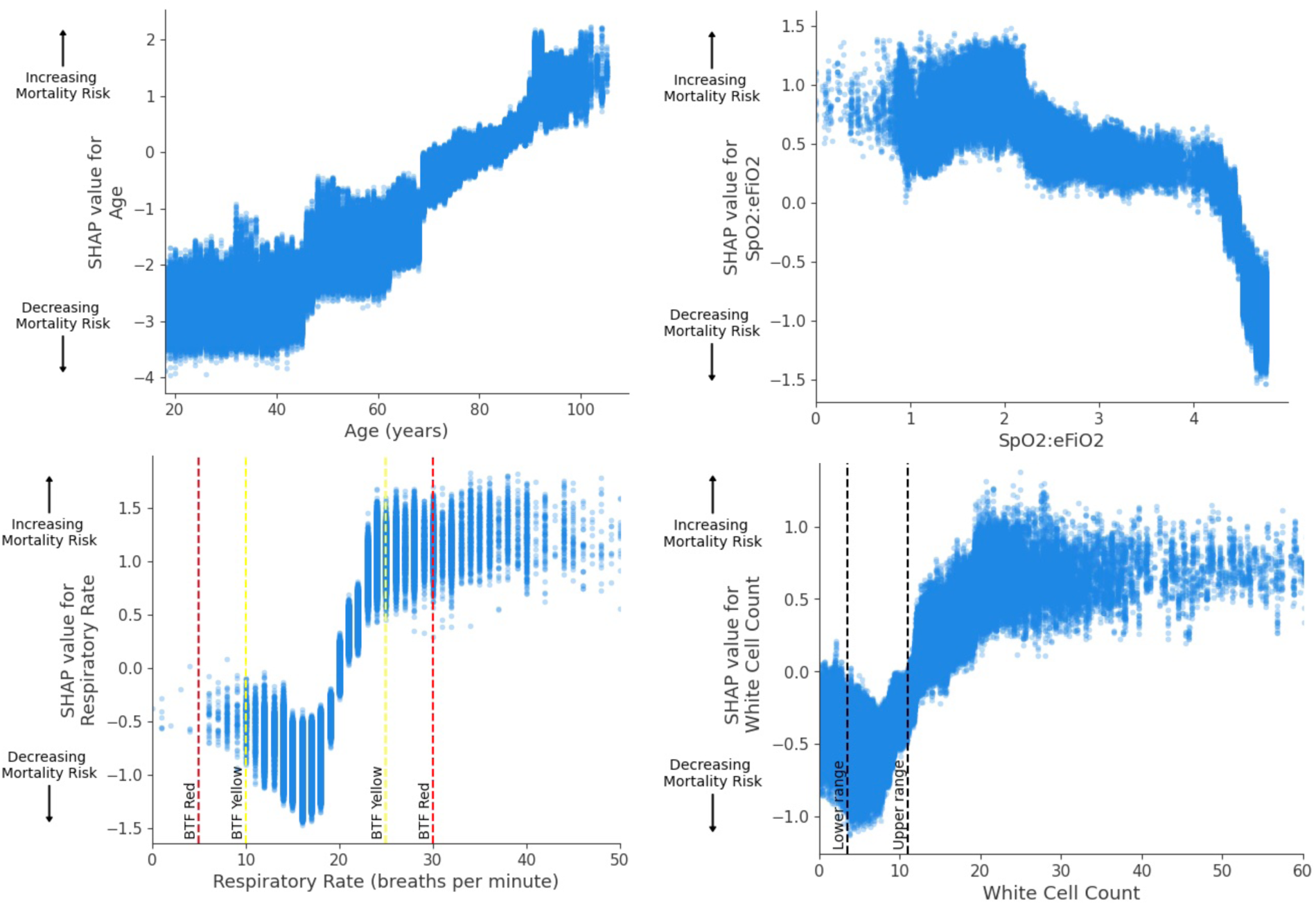
Partial dependence plots for 4 most influential predictors of mortality within 24 hours. Dashed lines indicate reference ranges, and colours indicate Between the Flags intervals where relevant. A positive value indicates an increase in the probability of the outcome. SpO2:eFiO2, ratio of oxygen saturation to estimated fraction of inspired oxygen.

**Figure 5:**
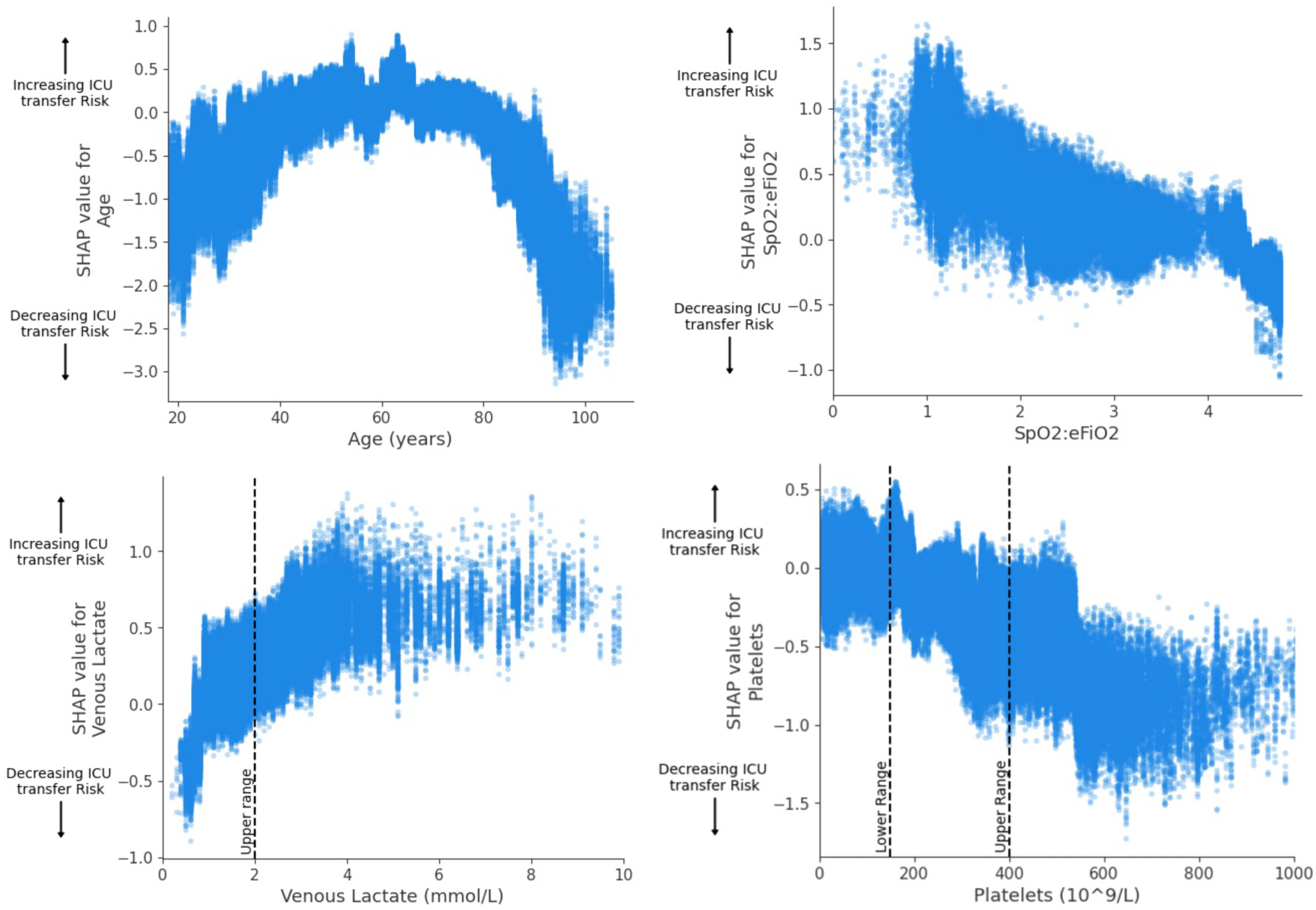
Partial dependence plots for 4 most influential predictors of unplanned ICU transfer within 24 hours. SpO2:eFiO2, ratio of oxygen saturation to estimated fraction of inspired oxygen; Shock Index, ratio of heart rate to systolic blood pressure. Dashed lines indicate reference ranges.

The Shapley partial dependence plot in Figure 6(a) explores the physiological interrelations of model predictions, where a low haemoglobin (blue) is correlated with stronger prediction of deterioration in scenarios of hypotension; potentially reflecting, for instance, a more acute pathology (i.e. bleed). The partial dependence plot for heart rate correlated with age in Figure 6(b) shows that age modulates the severity of an abnormal heart rate measurement. A slow heart rate is weighted more heavily towards predicting ICU transfer in an older patient than in a younger patient, whereas a faster heart rate (>100bpm) in younger patients indicates an increased risk of an ICU transfer.

**Figure 6.**
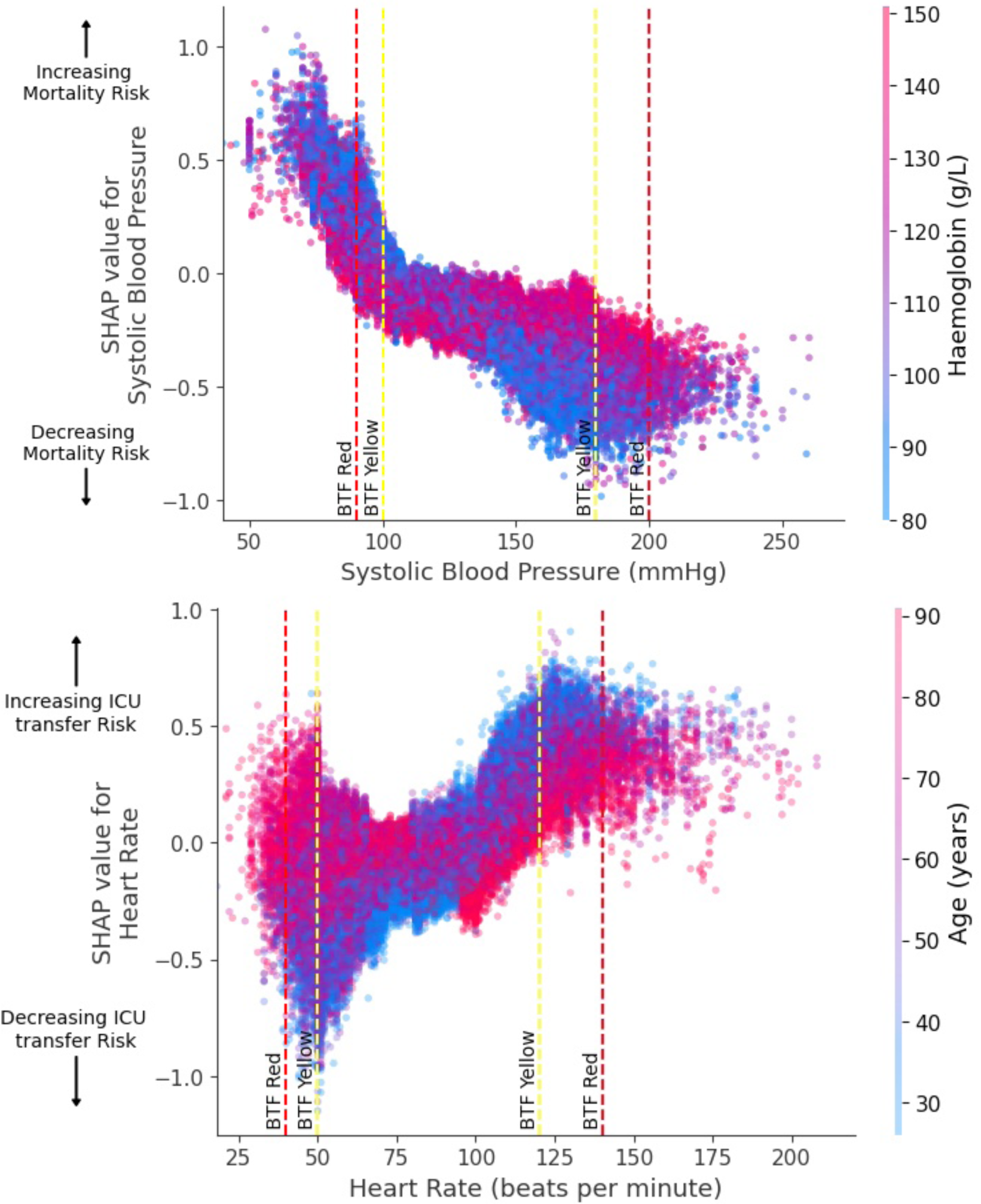
(a)(top): Partial dependence plot of SHAP values for systolic blood pressure predicting mortality with associated haemoglobin. Figure 6(b)(bottom): Partial dependence plot of SHAP values for heart rate predicting ICU transfer with associated age. Figures visualise the impact of the specified vital sign on model predictions, with higher SHAP values indicating higher relative weighting of the feature towards predicting the outcome of deterioration, and negative values weighting towards a negative prediction (no deterioration). The colour gradient represents the interacting lab value, showing how the interaction between the vital sign and lab value influences the model’s prediction of mortality. Vertical dashed lines indicate the colour coded Between the Flags thresholds for the corresponding vital sign

Shapley force plots (Figure 7) sum the relative positive and negative influence of features on a single point prediction and offers further depth to correlating model output with clinical values. Figure 7(a) explains a scenario where an elderly patient admitted under the Geriatric service experiences an isolated hypotension of 86mmHg (BTF red zone), with aWARE correctly weighting patient factors away from this heralding a deterioration outcome (no mortality or ICU transfer in this admission). Figure 7(b) shows the converse, with a correct positive prediction of deterioration 12 hours from death, for a middle-aged Cardiothoracic patient with all BTF vital signs in the normal range.

**Figure 7.**
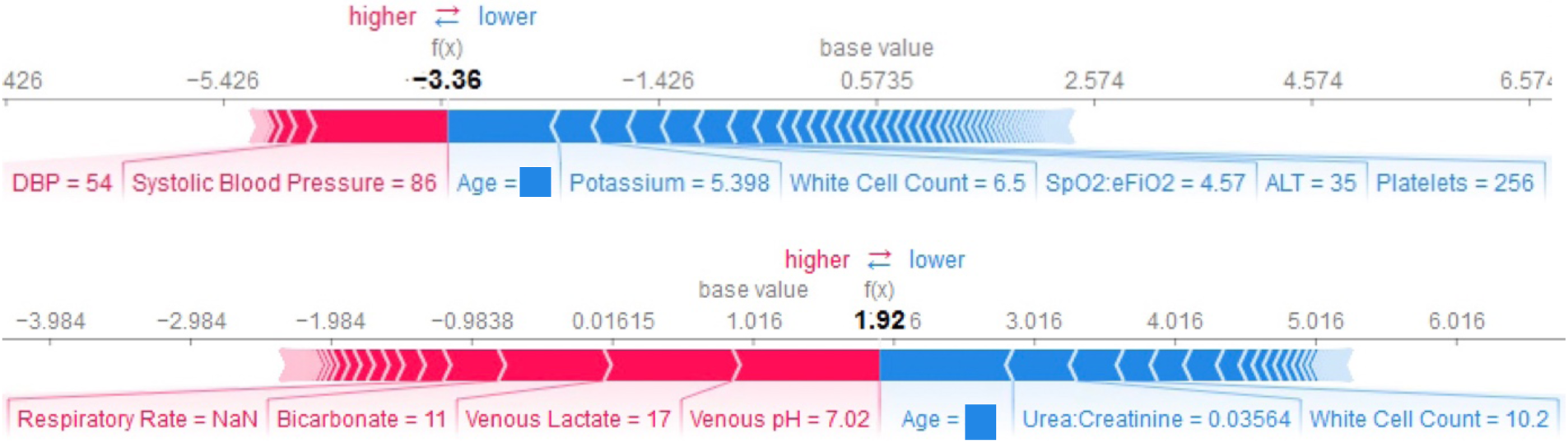
(a)(top): Force plot illustrating the Shapley values for a model negative prediction of ICU admission. Figure 7(b)(bottom): Force plot illustrating the Shapley values for a model positive prediction of mortality. The plots visualise the impact of individual features on the model’s prediction for a specific instance. Features pushing the prediction higher (towards deterioration) are shown in blue, while those pushing it lower (away from deterioration) are in red. Age has been omitted for deidentifying purposes. ALT, Alanine Transaminase; DBP, Diastolic blood pressure; SpO2:eFiO2, ratio of oxygen saturation to estimated fraction of inspired oxygen.

### Clinical Utility and Subgroup Analysis

Clinical utility (Figures S3 and S4) to summate impact and subgroup analysis (Tables S3 and S4) to examine for demographic bias are shown in the Supplementary Material.

## DISCUSSION

This study represents a promising application of a simple but highly efficient machine learning algorithm, Gradient Boosted Trees, for the identification of ward patients at risk of deterioration.

### Predictive Performance

BTF, MEWS and NEWS scores validated in the DREAM dataset were similar to prior studies^28^. aWARE outperformed all scoring systems internally tested on this dataset, including BTF. These results demonstrate that Gradient Boosted Trees are powerful for predicting deterioration, validating the work of previous similar models^17, 18^. To the authors’ knowledge, this study is the first to show similar applicability in the Australian hospital setting.

### Implications

The two aWARE models showed some divergence of influential predictors. This confirms that mortality and ICU transfer should be modelled as competing outcomes. Analysis of feature importance confirms that vital signs alone may not give a complete clinical picture of deterioration^17, 29, 30^. Enumerated in prior models, aWARE likewise increased in performance as demographics and pathology results, constructed variables, and linear trends were added beyond vital signs only^31, 32^. Additionally, aWARE’s ability to make predictions automatically even in the absence of pathology results, may broaden clinical implementation.

Partial dependence plots (Figures 6(a,b)) demonstrate two examples of how the various interactions between deranged patient variables which trigger MET calls (e.g. hypotension or bradycardia) may account for the false positives of simpler scores^33^. Depending on the value of certain laboratory values (e.g. haemoglobin) or demographic baseline characteristics (e.g. age), a single abnormal vital sign (e.g. systolic blood pressure or heart rate) shows variance in its weighted effect towards predicting deterioration, where the absolute thresholds of BTF or MEWS/NEWS do not. The addition of linear trends incorporated into this model stratify the risk of acutely against chronically deranged patient variables, enhancing model accuracy. The force plots (Figure 7(a,b)) show the potential for a model integrated within the eMR to display prediction rationale as needed, allowing the clinician to integrate their own judgement of the interplay of deteriorating patient factors.

Early detection and intervention of deteriorating patients has been shown to reduce adverse outcomes^34–36^. As aWARE displays an increased sensitivity to detect deterioration earlier than BTF, its implementation represents a strong opportunity to improve patient outcomes. Adequate warning of potential deterioration may also allow time for facilitated ceilings-of-care discussions, which may manifest as decreased unnecessary costly interventions^32^. Use of the aWARE model may yield economic benefit through reduction of false positive alerts and MET calls.

### Strengths and Limitations

The data used to train this model was adequately large to develop a machine learning model of deterioration. Despite this, given that risk of ICU transfer decreased in the oldest (>85) age cohort in Figure 5, the training data for predicting ICU transfer was evidently biased by the lack of palliation data needed to exclude older patients who were not indicated for ICU care. Where this model excluded patients admitted to the Palliative Care service, future iterations necessitate training datasets with more specific determination of end-of-life care for patient exclusion, such as Advanced Care Directives and Do Not Resuscitate orders. Additionally, local hospital policy and case mix determine the indication for escalation, especially considering the variation in capabilities of high-risk units (CCU and HDU). Exclusion of these from the outcome of ICU transfer may have misidentified some deteriorating patients.

Therefore, implementation into other local health districts may require expert review of whether transfer to these ought to be included in the primary outcome.

Comparing the developed model to the current standard of care by matching closest false positive rates allows for easy interpretation of the model’s superiority over BTF. This use case demonstrated clear gains in early detection without increasing false positives. A comparison to observed MET calls, not available in this data, rather than calculated BTF alerts, would provide a more accurate assessment of the model’s expected utility gain.

The included predictors were not exhaustive. Future models require expert review of variables and should explore the value of other predictors not currently included, such as inflammatory markers and arterial blood gas results. Predictions may be enhanced by multiclass prediction of specific outcomes of deterioration – including cardiac arrest, sepsis, acute kidney injury or need for mechanical ventilation^37, 38^. Furthermore, the addition of out-of-hospital outcomes may improve the accuracy of deterioration prediction.

While the model performed strongly on this dataset, it has only been internally validated. External validation trials are needed in hospitals to test the predictive performance and generalisability of the algorithm, and to measure patient outcomes to assess clinical benefit.

## Conclusion

There are currently no machine learning models that have been widely implemented in Australia which monitor patients and identify deterioration. This study proposed the aWARE method for prediction of in-hospital patient deterioration. This included extended analyses of feature importance, which highlight the deficit between the capabilities of an electronic deterioration tool which integrates large eMR data, and current practises of simple vital sign tracking. Ultimately, the performance of aWARE, developed for Australian health standards, demonstrates clear superiority to the outdated use of Between the Flags by identifying more at-risk patients while reducing false positive alerts. For the patient to reap the full benefit of such an algorithm, external validation is needed.

## Data Availability

All data produced in the present work are contained in the manuscript

## Authorship and acknowledgements

Jonathan Greenberg contributed to conceptualisation, analysis and manuscript writing. Blanca Gallego was involved in conceptualisation, data curation and manuscript review. Leo Huberts assisted in conceptualisation and methodology and provided manuscript review. The DREAM database is a research-ready data repository of eMR from Sydney Local Health District (SLHD), developed as a collaboration between the SLHD and the University of New South Wales (UNSW). Angus Ritchie is the clinical lead of the DREAM project. Angus Ritchie, Sze-Yuan Ooi, Gordon Flynn and Graeme Hart provided clinical expertise and reviewed the manuscript. This research has been partly supported by the NSW Agency for Clinical Innovation (ACI) Grant Scheme 23/24. Its contents are the responsibility of the authors and their institutions and do not necessarily reflect the views of the ACI.

