## Supplementary Material for "Improved Sensitivity For Detection Of Clinical Deterioration When Diagnostic Pathology And Patient Trends Are Included In Machine Learning Models"

**Figure S1:**

Flowchart of study design and cohort sample size, with exclusion criteria.
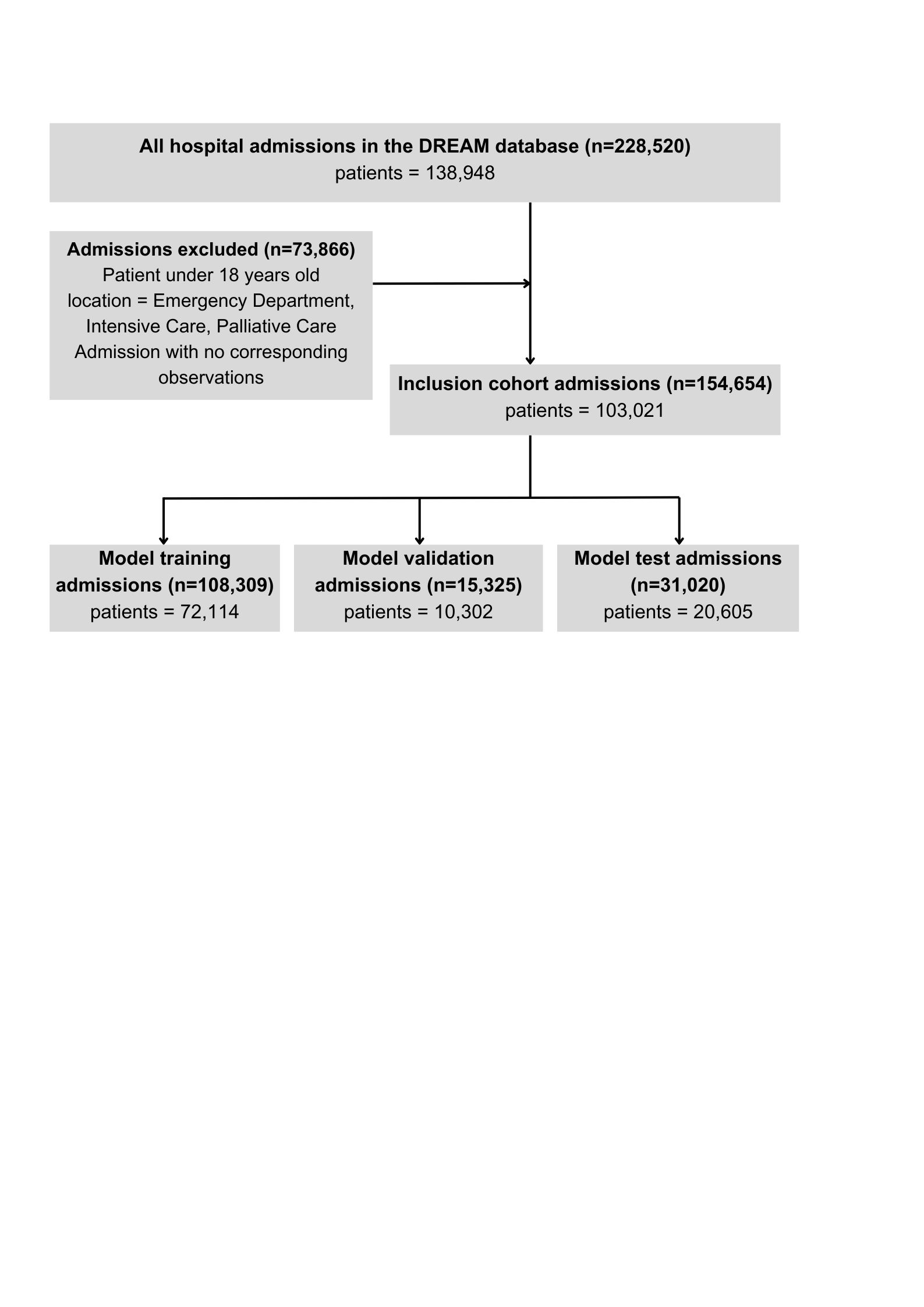

**Table S1: Demographic statistics of patients split by primary outcome**

| **Characteristic** | **Patients who did not die in-hospital in the study period** | **Patients who died in-hospital within the study period** |
| --- | --- | --- |
| Number of unique patients (% total patients) | 101,698 (98.72%) | 1,503 (1.28%) |
| Number of unique admissions (% total admissions) | 153,148 (99.12%) | 1,506 (0.88%) |
| Age (years)  Median  Interquartile Range (Q1 – Q3) | 67  (47 – 81) | 83  (74 – 89) |
| Gender  % male  % female | 45.9  54.1 | 56.8  43.2 |
| **Characteristic** | **Patients who were not transferred to ICU unplanned from hospital wards in the study period** | **Patients who were transferred to ICU unplanned from hospital wards in the study period** |
| Number of unique patients (% total patients) | 100,906 (97.95%) | 2,115 (2.05%) |
| Number of unique admissions (% total admissions) | 152,392 (98.53%) | 2,262 (1.46%) |
| Age (years)  Median  Interquartile Range (Q1 – Q3) | 68  (48 – 81) | 66  (55 – 76) |
| Gender  % male  % female | 46.1  53.9 | 58.9  41.1 |

**Table S2:**

All predictors used to generate an output in the algorithm.

| **Static patient variables** | **Dynamic patient variables** | | |
| --- | --- | --- | --- |
| **Demographics** | **Vital Signs** | **Laboratory Results** | **Trends** |
| Age  Gender | Heart rate  Respiratory rate  Systolic Blood Pressure  Diastolic Blood Pressure  Oxygen Saturation (SpO2)  Estimated Fraction of Inspired Oxygen (eFiO2)  Temperature  SpO2:eFiO2 **(constructed)**  Shock Index (Heart Rate:Systolic Blood Pressure) **(constructed)** | ALP  ALT  Albumin  Bicarbonate  Bilirubin Total  Calcium  Chloride  Creatinine  Estimated GFR  Glucose  Haemoglobin  Haematocrit  Platelets  Potassium  Red Cell Count  Sodium  Urea  Venous Base Excess  Venous Lactate  Venous pH  White Cell Count  Urea:Creatinine **(constructed)** | Baseline Trend for all vital signs and laboratory results  Trend from previous measurement for all vital signs and laboratory results  Trend from measurement two measurements prior for all vital signs and laboratory results |

**Figure S2: Cumulative percent of patients who died in hospital with any alerted at listed time intervals 48 hours preceding death. Figure S8: Cumulative percent of patients who were transferred to ICU unplanned from wards with any alerted at listed time intervals 48 hours preceding unplanned ICU transfer.**

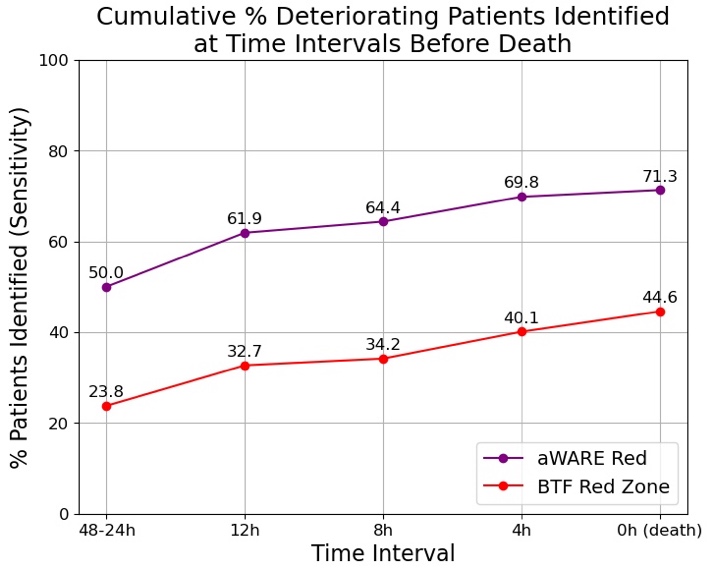
The cumulative percent of patients identified (sensitivity) with at least 1 BTF Red alert or aWARE Red alert at several time intervals in the 48 hours preceding the primary outcome time-point is shown in Fig. 5. At a patient level, aWARE identified more deteriorating patients than BTF Red Zone at all points leading to death or ICU transfer. Both BTF Red alerts and aWARE Red alerts increase in sensitivity closer to the event horizon. aWARE Red, ai-driven WArning and REsponse system matched to closest specificity of Between the Flags Red Zone; BTF Red Zone, Between the Flags red zone calling criteria

**Figure S3: Cumulative percent of patients who were transferred to ICU unplanned from wards with any alerted at listed time intervals 48 hours preceding unplanned ICU transfer.**

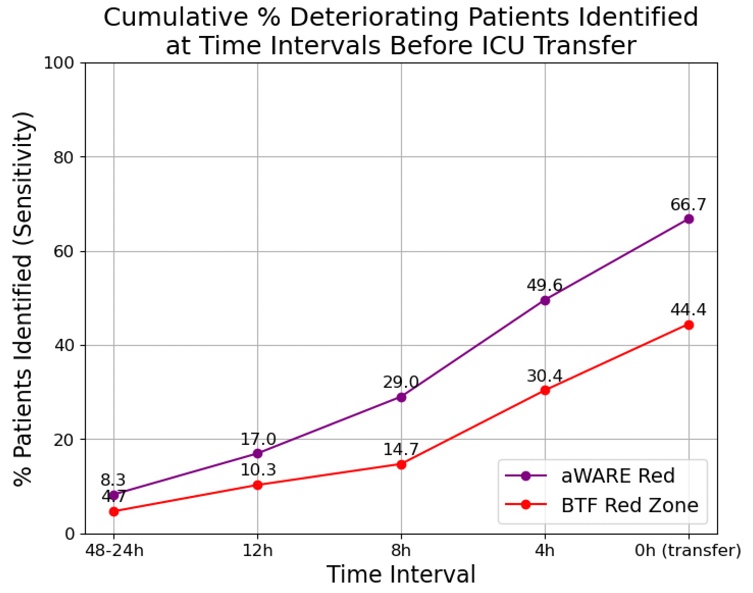
The cumulative percent of patients identified (sensitivity) with at least 1 BTF Red alert or aWARE Red alert at several time intervals in the 48 hours preceding the primary outcome time-point is shown. At a patient level, aWARE identified more deteriorating patients than BTF Red Zone at all points leading to death or ICU transfer. Both BTF Red alerts and aWARE Red alerts increase in sensitivity closer to the event horizon. aWARE Red, ai-driven WArning and REsponse system matched to closest specificity of Between the Flags Red Zone; BTF Red Zone, Between the Flags red zone calling criteria

**Figure S4: SHAP Beeswarm Summary plot for predicting mortality within 24 hours**

SHAP Value Beeswarm summary plot, showing the direction of prediction influence as a function of feature size. Positive SHAP values correspond to a greater influence toward a prediction of 1 (mortality <24h), while negative SHAP values correspond to a greater influence toward a prediction of 0 (no mortality <24h). Red points represent higher feature values (e.g. greater age) while blue represents lower feature values (e.g. lower respiratory rate). SpO2:eFiO2, ratio of oxygen saturation to estimated fraction of inspired oxygen; Shock Index, ratio of heart rate to systolic blood pressure; SpO2, Oxygen saturation.

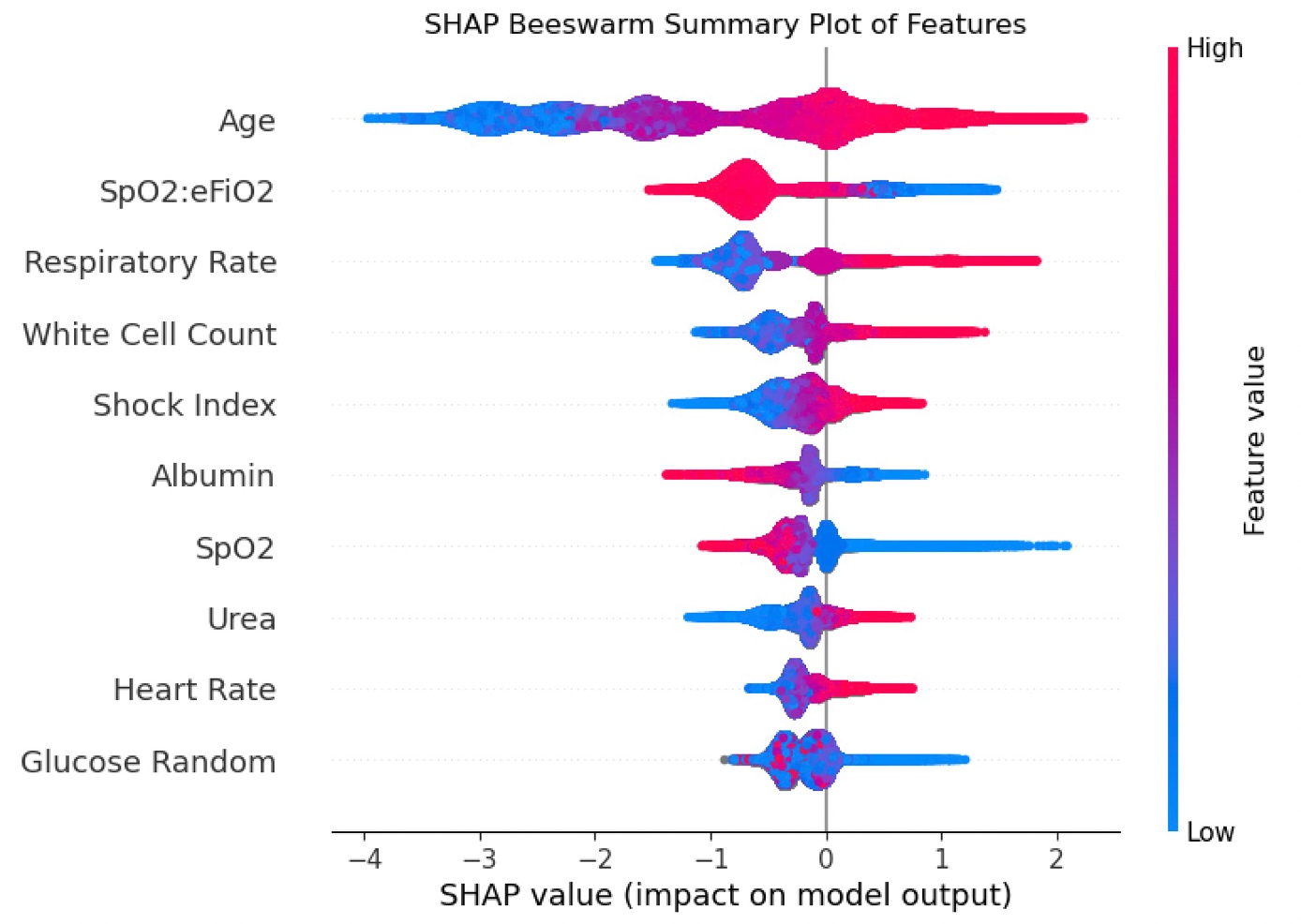

**Figure S5: SHAP Beeswarm Summary plot for predicting unplanned ICU transfer within 24 hours**

SHAP Value Beeswarm summary plot, showing the direction of prediction influence as a function of feature size. Positive SHAP values correspond to a greater influence toward a prediction of 1 (mortality <24h), while negative SHAP values correspond to a greater influence toward a prediction of 0 (no mortality <24h). Red points represent higher feature values while blue represents lower feature values. SpO2:eFiO2, ratio of oxygen saturation to estimated fraction of inspired oxygen; Shock Index, ratio of heart rate to systolic blood pressure.

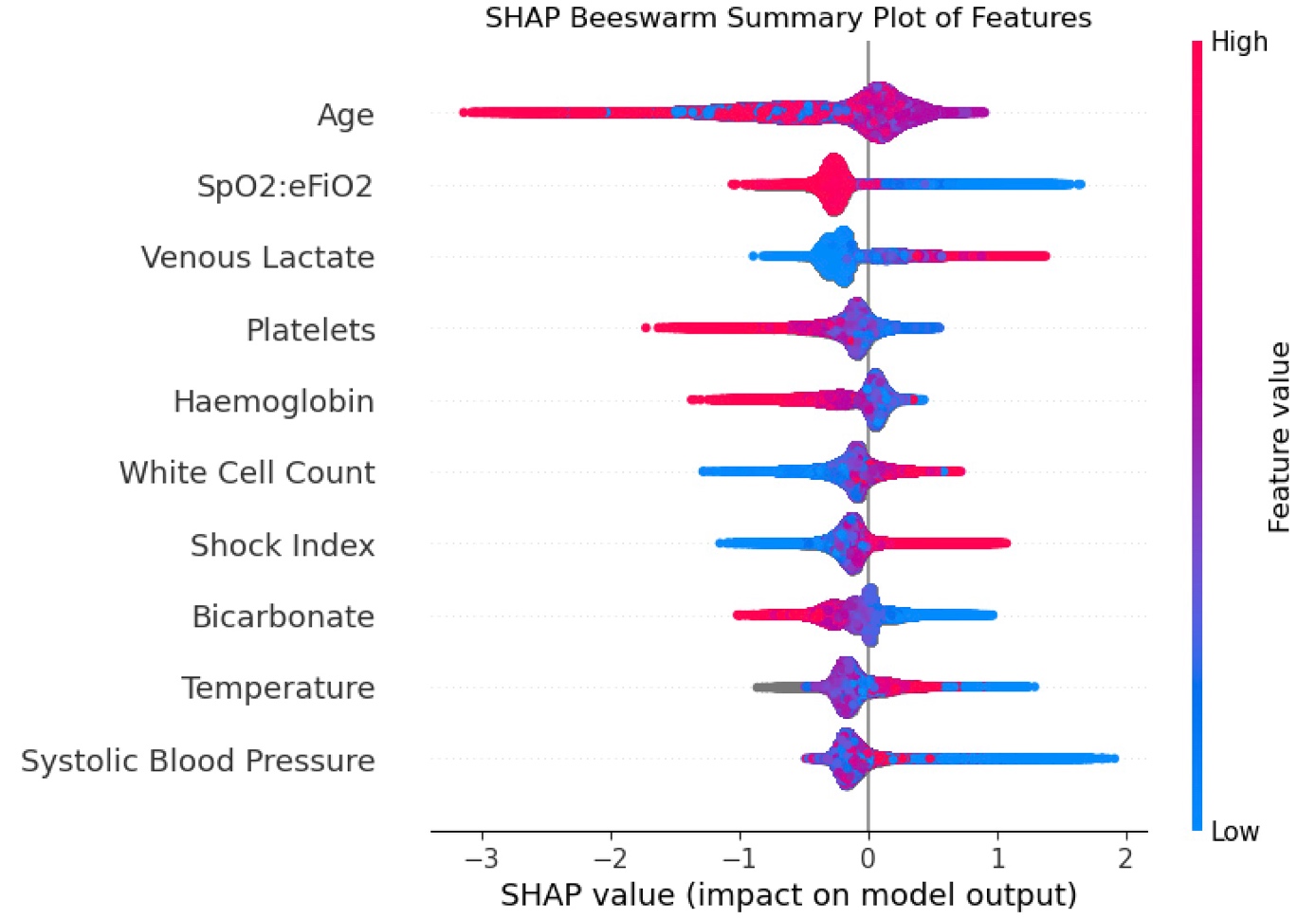

### **Figures S6: Expected Clinical Utility of aWARE predicting mortality**

### Net benefit curve is used in decision theory to evaluate the expected utility of predictive models by balancing the true positive rate (TPR) against the false positive rate (FPR), weighted by their respective benefits and harms(39). It is calculated as:

$$Net Benefit=p\cdot TPR\cdot B-(1-p)\cdot FPR\cdot C$$

Here, p is the event outcome prevalence, B is the expected net benefit of appropriate treatment associated with a true positive, and C is the expected net cost of unnecessary treatment associated with of a false positive. TPR corresponds to the True Positive Rate and FPR corresponds to the False Positive Rate of the algorithm. It has been assumed that no action takes place in the absence of an alarm. The ideal probability threshold at which an alarm is raised corresponds to the case in which the expected utility of never treating equals the expected utility of always treating. It reflects the probability threshold's implicit cost-benefit trade-off and is related to the C/B ratio:

$$\frac{C}{B}=\frac{p_{threshold}}{1- p_{threshold}}$$

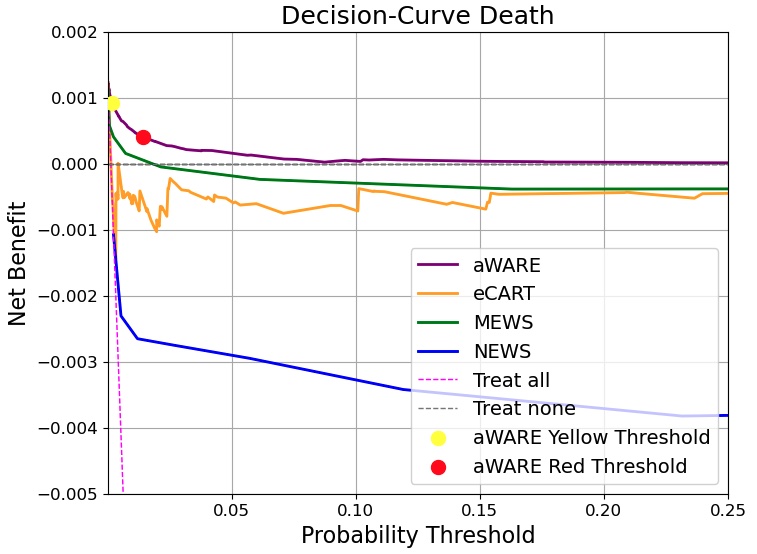
Figures show the net benefit curves of available clinical deterioration algorithms across different thresholds, indicating the expected clinical utility of models.

Decision curve analysis plots the varying model threshold against the Net Benefit. Curves are compared against strategies of Treat All and Treat None. Points are shown for the Net Benefit at the thresholds which match aWARE specificity to that of BTF.

The estimated Net Benefit values are very small, since the rate of deterioration per newly available measurement (p_death_=0.140%, p_ICU transfer_=0.722% ) is extremely small. Nevertheless, aWARE displays a higher net benefit curve than eCART, MEWS and NEWS for all thresholds.

aWARE, ai-drive WArning and REsponse system; BTF, Between the Flags. MEWS, Modified Early Warning Score; NEWS, National Early Warning Score

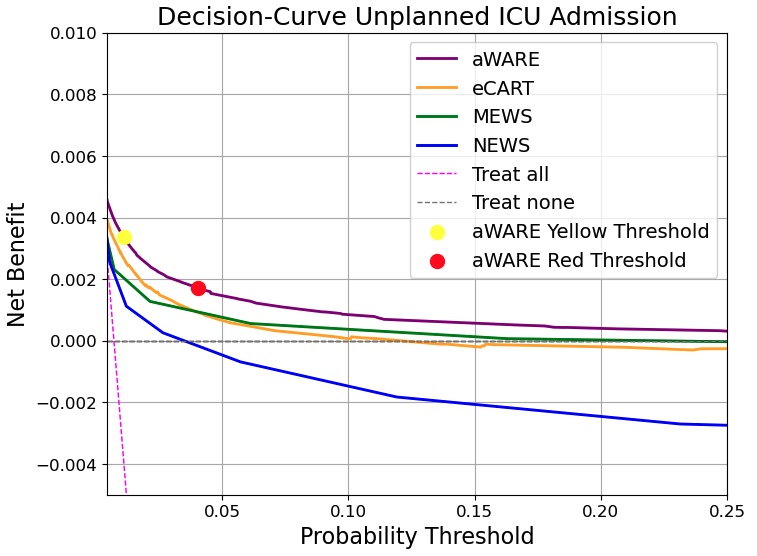

### **Figure S7: Expected Clinical Utility of aWARE predicting ICU admission**

aWARE, ai-drive WArning and REsponse system; BTF, Between the Flags. MEWS, Modified Early Warning Score; NEWS, National Early Warning Score

**Table S3: Subgroup Analysis for prediction of death within 24 hours**

Only the 10 services with the most frequent number of deterioration of events (observations within 24h of death) are shown.

aWARE, aWARE, ai-driven WArning and Response system; BTF, Between the Flags; TPR, TruePositive Rate; TNR, True Negative Rate.

|  | **aWARE**  **Red**  **TPR** | **aWARE**  **Red**  **TNR** | **BTF Red TPR** | **BTF Red TNR** | **BTF Yellow TPR** | **BTF Yellow TNR** | **Event Number** | **Event Rate %** |
| --- | --- | --- | --- | --- | --- | --- | --- | --- |
| **Gender** |  |  |  |  |  |  |  |  |
| Female | 0.583 | 0.983 | 0.294 | 0.976 | 0.278 | 0.873 | 544 | 0.090 |
| Male | 0.467 | 0.975 | 0.229 | 0.981 | 0.260 | 0.889 | 1083 | 0.195 |
| **Age group** |  |  |  |  |  |  |  |  |
| <45 | 0.263 | 1.000 | 0.000 | 0.977 | 0.237 | 0.844 | 38 | 0.015 |
| 45-54 | 0.111 | 0.995 | 0.156 | 0.978 | 0.289 | 0.875 | 45 | 0.042 |
| 55-64 | 0.426 | 0.996 | 0.080 | 0.984 | 0.176 | 0.889 | 176 | 0.112 |
| 65-74 | 0.427 | 0.985 | 0.178 | 0.980 | 0.249 | 0.895 | 225 | 0.112 |
| 75-84 | 0.348 | 0.968 | 0.323 | 0.977 | 0.264 | 0.894 | 387 | 0.164 |
| <85 | 0.664 | 0.938 | 0.294 | 0.975 | 0.294 | 0.895 | 756 | 0.374 |
| **Service** |  |  |  |  |  |  |  |  |
| Geriatric Medicine | 0.590 | 0.955 | 0.354 | 0.981 | 0.252 | 0.908 | 556 | 0.288 |
| Medicine -General | 0.677 | 0.970 | 0.257 | 0.989 | 0.181 | 0.935 | 210 | 0.192 |
| Cardiology | 0.447 | 0.967 | 0.244 | 0.942 | 0.479 | 0.812 | 163 | 0.175 |
| Haematology | 0.211 | 0.984 | 0.151 | 0.975 | 0.349 | 0.842 | 152 | 0.280 |
| Cardiothoracic Surgery | 0.500 | 0.963 | 0.025 | 0.981 | 0.400 | 0.875 | 120 | 0.344 |
| Gastroenterology | 0.550 | 0.991 | 0.300 | 0.975 | 0.280 | 0.875 | 100 | 0.270 |
| Renal Medicine | 0.303 | 0.985 | 0.180 | 0.968 | 0.236 | 0.881 | 89 | 0.291 |
| Neurosurgery | 0.802 | 0.978 | 0.058 | 0.986 | 0.070 | 0.908 | 86 | 0.354 |
| Thoracic Medicine | 0.167 | 0.933 | 0.053 | 0.956 | 0.154 | 0.857 | 78 | 0.211 |
| Orthopaedics | 0.500 | 0.992 | 0.045 | 0.989 | 0.000 | 0.904 | 22 | 0.031 |

**Table S4: Subgroup Analysis for prediction of ICU transfer within 24 hours**

Only the 10 services with the most frequent number of deterioration of events (observations within 24h of ICU transfer) are shown.

aWARE, aWARE, ai-driven WArning and Response system; BTF, Between the Flags; TPR, True Positive Rate; NR, True Negative Rate.

|  | **aWARE**  **Red**  **TPR** | **aWARE**  **Red**  **TNR** | **BTF Red TPR** | **BTF Red TNR** | **BTF Yellow TPR** | **BTF Yellow TNR** | **Event Number** | **Event Rate** |
| --- | --- | --- | --- | --- | --- | --- | --- | --- |
| **Gender** |  |  |  |  |  |  |  |  |
| Female | 0.561 | 0.927 | 0.229 | 0.977 | 0.297 | 0.874 | 3431 | 0.565 |
| Male | 0.646 | 0.874 | 0.241 | 0.982 | 0.275 | 0.890 | 4961 | 0.894 |
| **Age group** |  |  |  |  |  |  |  |  |
| <45 | 0.487 | 0.957 | 0.277 | 0.978 | 0.291 | 0.845 | 1317 | 0.509 |
| 45-54 | 0.667 | 0.860 | 0.223 | 0.980 | 0.293 | 0.877 | 1002 | 0.937 |
| 55-64 | 0.621 | 0.849 | 0.269 | 0.986 | 0.249 | 0.890 | 1511 | 0.963 |
| 65-74 | 0.700 | 0.861 | 0.225 | 0.982 | 0.296 | 0.896 | 1921 | 0.954 |
| 75-84 | 0.631 | 0.890 | 0.220 | 0.978 | 0.305 | 0.896 | 1716 | 0.726 |
| <85 | 0.494 | 0.947 | 0.194 | 0.975 | 0.256 | 0.895 | 925 | 0.457 |
| **Service** |  |  |  |  |  |  |  |  |
| Upper GIT | 0.600 | 0.875 | 0.173 | 0.992 | 0.219 | 0.900 | 813 | 1.951 |
| Thoracic Medicine | 0.775 | 0.820 | 0.427 | 0.964 | 0.283 | 0.860 | 757 | 2.046 |
| Colorectal Surgery | 0.426 | 0.888 | 0.050 | 0.991 | 0.124 | 0.889 | 659 | 1.434 |
| Cardiology | 0.596 | 0.846 | 0.309 | 0.944 | 0.280 | 0.812 | 653 | 0.701 |
| Haematology | 0.601 | 0.904 | 0.345 | 0.978 | 0.424 | 0.844 | 589 | 1.086 |
| Renal Medicine | 0.645 | 0.865 | 0.248 | 0.972 | 0.339 | 0.884 | 561 | 1.833 |
| Gastroenterology | 0.777 | 0.878 | 0.286 | 0.978 | 0.357 | 0.860 | 538 | 1.450 |
| Geriatric Medicine | 0.649 | 0.951 | 0.317 | 0.981 | 0.304 | 0.908 | 527 | 0.273 |
| Vascular Surgery | 0.656 | 0.881 | 0.040 | 0.990 | 0.220 | 0.897 | 378 | 1.644 |
| Orthopaedics | 0.417 | 0.888 | 0.138 | 0.991 | 0.274 | 0.904 | 369 | 0.519 |
